# Predicting COVID-19 incidence from seroprevalence and population-based cohort data using interpretable machine learning with differential privacy analysis

**DOI:** 10.64898/2026.04.01.26349876

**Authors:** Jessica Krepel, Ruta Binkyte, Raouf Kerkouche, Manuela Harries, Carolina J. Klett-Tammen, Mario Fritz, Stefan Kesselheim, Martin Kühn, Alina Bazarova, Berit Lange, with the MuSPAD study group, RESPINOW study group

## Abstract

During the COVID-19 pandemic, reported incidence data played a central role in public health surveillance and in tracking epidemic dynamics, although they provide limited insight into the behavioral, immunological, and socioeconomic drivers of transmission.Population-based seroprevalence studies with linked survey data offer a rich but untapped source of individual-level information that can complement routine surveillance. In this study, we investigate whether aggregated seroprevalence cohort data can be leveraged to predict local COVID-19 incidence and to identify interpretable predictors associated with transmission dynamics. Using data from the Multilocal SeroPrevalence (MuSPAD) study in Germany (2020–2022), we trained multiple machine learning models, including least absolute shrinkage and selection operator (LASSO), vector autoregressive models (VAR), multilayer perceptrons (MLPs), and long short-term memory neural networks (LSTMs), to predict location-specific seven-day incidence rates. Feature importance was assessed using regression coefficients where applicable and model-agnostic explainability methods, including Local Interpretable Model-agnostic Explanations (LIME) and SHapley Additive exPlanations (SHAP). Across model classes, cohort-derived features enabled accurate prediction of local incidence, with time-aware models achieving the strongest performance. Consistent predictors included prior infection and testing history, employment-related changes, vaccination status, and mask-wearing behavior, highlighting the importance of behavioral and reporting-related signals. While differential privacy introduced modest degradation in predictive performance under strict privacy budgets, SHAP-based explanations remained stable, and LIME-based explanations were more sensitive to privacy-induced noise. These results demonstrate that aggregated cohort data encode meaningful and interpretable signals of population-level transmission dynamics. Population-based serosurveys therefore provide a complementary source of information for predicting local COVID-19 incidence and identifying key drivers of transmission beyond routine surveillance data. Our findings show that integrating interpretable machine learning with privacy-aware analysis enables actionable insights from sensitive cohort data, supporting their use in digital epidemiology and informing data-driven public health decision-making.

**Author summary:** During the COVID-19 pandemic, public health decisions were largely based on reported case numbers. However, these data provide limited insight into the behavioral and social factors that influence how infections spread. Population-based studies that combine antibody testing with questionnaires can capture such information, but they are rarely used to predict disease dynamics at the population level. In this study, we investigated whether data from a large seroprevalence cohort study in Germany could be used to predict local COVID-19 incidence. We applied machine learning models to aggregated survey and laboratory data and used explainability methods to identify which factors were most important for the predictions. We also examined how protecting individual privacy affects both model performance and interpretation. We found that cohort-derived features can accurately predict short-term incidence trends. Key factors included prior infections, testing behavior, employment-related changes, vaccination status, and mask use. While strong privacy constraints reduced model performance, the main insights remained stable. Our results show that combining cohort data with interpretable machine learning can provide useful insights into disease dynamics. This approach may help improve future public health surveillance by incorporating information that is not captured in routine reporting systems.

## 1 Introduction

The COVID-19 pandemic caused by severe acute respiratory syndrome coronavirus 2 (SARS-CoV-2) has had an unprecedented impact on global health and society, prompting the need for accurate and timely data on disease spread, immunity, and risk factors. Between 2020 and 2022, the World Health Organization reported more than 660 million confirmed cases and nearly 7 million deaths worldwide [1], placing the forecasting of disease dynamics at the center of public health decision-making. Throughout the pandemic, governments implemented a range of control policies that evolved over time in response to emerging scientific evidence, shifting epidemiological trends, and societal needs. Consequently, numerous modeling approaches and data sources have been explored to predict disease incidence. These include mechanistic, hybrid, and data-driven machine learning (ML) models leveraging diverse data types such as reported incidence [2–4], wastewater measurements [5–9], human mobility [3, 4, 10], and cell phone–derived data [11, 12]. In addition, seroprevalence studies have been used to inform mechanistic models [3, 13, 14].

In this study, we analyze data from the Multilocal SeroPrevalence (MuSPAD) study, a populationbased cross-sectional seroepidemiological cohort conducted in Germany during the COVID-19 pandemic. MuSPAD provides detailed individual-level data from a representative sample of adults, including serological measurements and questionnaire-based information on household structure, behavior, and exposure. Previous analyses have used these data to estimate infection frequency, underdetection, and infection fatality across multiple epidemic phases [15–19]. Through repeated sampling waves and integration of laboratory measurements, the study enables temporally resolved monitoring of infection dynamics and supports adaptive responses to emerging epidemiological questions. When combined with complementary data sources, such cohort data can provide a more comprehensive view of population-level disease dynamics [20, 21].

While machine learning (ML) methods have been widely applied to infectious disease prediction [22] and to the development of surrogate models for mechanistic approaches [23], their use with time-resolved, population-based cohort data remains comparatively limited, despite the broad adoption of ML across diverse modeling paradigms [24]. Systematic reviews of ML in infectious disease research document extensive work on surveillance, diagnosis, prognosis, and outbreak forecasting, but also highlight persistent challenges related to generalizability, validation, and transferability across cohorts and settings [25].

Existing ML applications in infectious disease modeling largely align the level of the prediction target with the level of the available data. Population-level incidence or spread is typically predicted from aggregated covariates, such as country-or region-level non-pharmaceutical interventions [22, 26]. Conversely, studies that leverage individual-level cohort data—including surveys and serological measurements—predominantly focus on person-level outcomes, such as clinical prognosis [27], individual infection or severity risk [28], or vaccination and infection status classification [29]. As a result, ML-based approaches that aggregate individual-level cohort information to infer population-level disease dynamics remain comparatively rare. A few notable exceptions include models using aggregated sociodemographic and survey-derived features to estimate neighborhood-level disease prevalence [30], or training ML algorithms on individual cohort and administrative health records to predict population-level incidence, such as diabetes in France [31].

In parallel, reviews of explainable AI (XAI) in health applications report widespread adoption of post hoc interpretation methods, such as SHAP and LIME, while highlighting limitations due to dataset heterogeneity and challenges in clinical or epidemiological applicability [32, 33]. Integrating ML with XAI to systematically identify key drivers of population-level disease dynamics in longitudinal seroepidemiological cohorts, such as MuSPAD, remains largely unexplored. This gap highlights the potential of such approaches to extract interpretable insights from rich individual-level data.

Here, leveraging the large number of participants, we investigate whether aggregated MuSPAD cohort data can be used to predict local COVID-19 incidence and to identify interpretable determinants of transmission using ML models. This approach also highlights relevant features that are typically absent from public health agency datasets. Interpreting the resulting predictive models is therefore essential not only for understanding and managing disease incidence, but also for informing the design of measurement systems and survey instruments in future epidemics to improve forecasting accuracy.

In addition, we extend our analysis to models trained under privacy constraints. Ensuring the privacy of individuals in ML applications is critical, particularly for sensitive health data, and is required for compliance with regulatory frameworks such as the General Data Protection Regulation (GDPR) and the Health Insurance Portability and Accountability Act (HIPAA), as well as for maintaining the trust of data contributors.

Prior studies have demonstrated that even when personal identifiers are removed, sensitive attributes can be inferred from model parameters or outputs, particularly when an attacker can combine the model with external auxiliary data [34]. To overcome these challenges, we employ the Differential Privacy (DP) framework, which is one of the most well-established methods. The core strength of DP is providing quantifiable, theoretically grounded privacy guarantees that hold under any transformations of the data, including ML. However, DP requires injecting calibrated noise during training, which can reduce model accuracy. This trade-off between privacy preservation and utility is particularly consequential in the medical domain, where both strong privacy protection and high predictive performance are essential. Hereby, we conduct a series of experiments across multiple privacy budgets to identify an optimal balance between accuracy and privacy for the MuSPAD data. In addition, we repeat the explainability analysis on the privatized data to assess the robustness of incidence-rate predictors to DP noise. In prior work [35], we have already demonstrated how DP can be used with federated learning on county-based confirmed cases for SARS-CoV-2.

This manuscript is structured as follows. Following the section 1, we describe the methodology, including data preparation, predictive models, baseline comparisons, and the explainability approaches employed, along with the application of differential privacy. Section 3 presents the study’s findings in tabular and graphical form, followed by section 4, which interprets these results in a broader context and concludes with a summary of key insights and implications.

## 2 Materials and methods

### 2.1 Data source and variables

We used data from the MuSPAD study comprising two successive cross-sectional surveys (2020– 2021), including over 32,000 participants across eight German regions. Each participant provided blood samples (collected using barcoded serum-gel Monovettes) and questionnaire responses. Follow-up data collected after 2021 were excluded due to substantial changes in questionnaire content. Daily seven-day COVID-19 incidence rates per 100,000, stratified by county, were obtained from the Robert Koch-Institute [36] and used as labels for the ML models.

To reduce dimensionality and noise, duplicate and low-informative variables were removed, reducing the feature space from 704 to 77 variables (see Supplementary Data). The retained variables captured factors expected to vary at the population level during the pandemic (e.g., employment-related changes), whereas largely time-invariant individual characteristics (e.g., date of birth) were excluded.

### 2.2 Data pre-processing of MuSPAD variables

In order to aggregate the individual-level data to population level, records from all participants attending on the same day were combined as follows. For numeric variables, the mean of all values was calculated. For categorical and binary variables *P*, the occurrence of each category *C*_1_, *C*_2_, …, *C*_*n*_ for every time point *t* was encoded into separate variables *Y*_1_, *Y*_2_, …, *Y*_*n*_ with

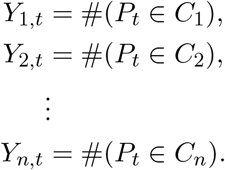

Date variables were converted to numeric values by subtracting January 1, 2020 from the participation date. We distinguished between explicitly encoded missingness (e.g., PCR info missing), which was retained as an informative feature, and true missing values (NA entries). Antibody test results and vaccination dates with more than 20% missing values were excluded to avoid introducing additional assumptions through imputation under substantial missingness, which could have biased the estimates. Variables corresponding to rare categories (e.g., responses such as “I do not know my test result”), which exhibited near-zero variance, were also removed, as they contributed negligible information to the analysis.

The resulting MuSPAD dataset comprised 279 observation days and 122 variables. This increase from the original 74 variables is due to the expansion of categorical variables into separate indicator variables during aggregation.

### 2.3 Model types and dataset construction

We evaluated two categories of models, distinguished by how they handle temporal structure. Time-agnostic models treat each day’s features independently, while time-aware models explicitly leverage temporal dependencies in the incidence data. For both model types, the input consisted of preprocessed MuSPAD variables combined with daily seven-day COVID-19 incidence per region from the Robert Koch-Institute [37].

For time-agnostic models, each data point represented feature values for a specific day, with the corresponding incidence seven days later as the prediction target. We applied regularized regression (LASSO) [38] and a multilayer perceptron (MLP) [39] to this dataset, which contained 279 daily observations and 123 variables (122 features and 1 target). The first 90% of days formed the training–validation set, and the remaining 10% the test set. Missing values were imputed separately in training–validation and test sets using the random-forest-based algorithm missForest [40]. Z-score normalization was computed on the training set and applied to the validation and test sets. For MLPs, the training–validation set was further split, with 10% used as a validation subset for early stopping and model selection. The final model was retrained on the full training–validation set prior to testing.

For time-aware models, including vector autoregressive (VAR) models [41] and long short-term memory (LSTM) networks [42], we incorporated past disease incidence as features alongside MuSPAD variables. Days without sampling were added with available incidence values and missing features, which were subsequently imputed. The resulting dataset contained 408 daily samples with 123 variables. For the seven-day prediction task, the most recent seven days were used as the test set, the preceding seven days as the validation set, and all earlier days as training data. To assess robustness and increase variation in test-set incidence values, models were also trained on the first three-quarters of the time series and evaluated on the subsequent seven days. Imputation and scaling followed the same procedure as for the MLP models.

This framework ensured consistent treatment of missing data, normalization, and temporal structure, providing a comparable basis for evaluating both time-agnostic and time-aware models, with test sets reflecting the most recent observations in the study period.

### 2.4 Machine learning methods

#### 2.4.1 LASSO regression

We performed LASSO regression defined as follows

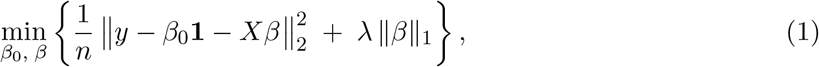

where *y ∈* ℝ ^*n*^ is the incidence variable, *X ∈* ℝ ^*n×p*^ is a design matrix, *β ∈* ℝ ^*p*^ is a vector of coefficients with *β*_0_ being an intercept, and *λ* is a penalty term.

We implemented LASSO regression using the R software environment [43] and the `glmnet` package [44], selected for its transparent and interpretable variable-selection properties. MuSPAD variables were used as predictors, with values observed on day *t* employed to predict the seven-day regional COVID-19 incidence on day *t* + 7.

During validation, 10-fold cross-validation on the training–validation set was used to select the regularization parameter *λ* minimizing the root mean squared error (RMSE). The final LASSO model was then retrained on the full training–validation set and evaluated on the test set.

#### 2.4.2 Multilayer perceptron

We compared LASSO results with those of a fully connected multilayer perceptron (MLP). The analysis was performed in R using the *tensorflow* [45] and *keras* [46] libraries. To avoid overparameterization given the limited number of datapoints, we evaluated architectures with one to four hidden layers and varied key hyperparameters during validation. Models were trained for 30, 45, and 60 epochs using both Adam and RMSprop optimizers. The final model, selected based on validation performance, utilized the Adam optimizer, was trained for 30 epochs with a batch size of 10% of the training data, and consisted of four hidden layers with 10 neurons each and ReLU activation. The output layer used a linear activation function for regression, with mean squared error (MSE) as the loss function.

#### 2.4.3 Vector auto-regression (VAR)

We performed a sparse estimation of a VAR model [41] to incorporate temporal dynamics while maintaining regularization through hierarchical lagged structures. This approach corresponds to group LASSO regularization with nested groups, such that

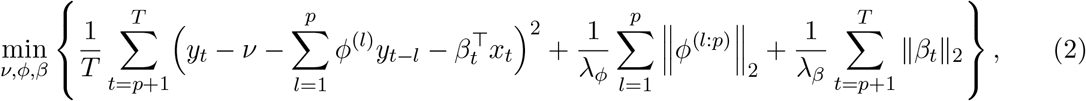

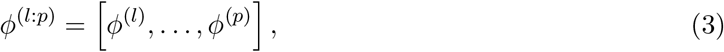

where *y*_*t*_ *∈* ℝ is the incidence value at time *t, v ∈* ℝ is the intercept (constant) term, *ϕ*^(*l*)^ *∈ ℝ* is the coefficient for the *l*-th lag (for *l* = 1, …, *p*). Each *ϕ*^(*l*)^ multiplies the lagged variable *y*_*t™l*_, *p* is the maximum lag order of the VAR model, *x*_*t*_ *∈ℝ* ^*m*^ is a vector of features, and *β*_*t*_ *∈ ℝ*^*m*^ is a vector of time-varying coefficients. *T* is the number of time–period observations (*t* = 1, 2, …, *T*), and *λ*_*ϕ*_ and *λ*_*β*_ are the respective penalty terms. We used the R-package *bigtime* [47] to fit a sparse VAR model and generate seven-day forecasts. Specifically, we estimated the model using `sparseVAR` with a penalization parameter set to either `HLag` (hierarchical penalty) or `L1` (standard LASSO) and explored the `selection` criterion among `cv` (time-series cross-validation), `bic` and `aic` corresponding to information criteria `AIC` and `BIC` which are supposed to balance model fit and complexity by penalizing the likelihood according to the number of parameters. We varied `cvcut` between 0.8 and 0.9, setting forecast horizon *h* to seven days. After model estimation, we used `recursiveforecast` to generate out-of-sample predictions on the test set. The best model, selected via validation, employed an *L*_1_ (LASSO) penalty with time-series cross-validation, and the optimal regularization parameters *λ*_*ϕ,β*_ were determined using 10-fold cross-validation, analogous to the LASSO procedure from Section 2.4.1. We assessed the models with different numbers of lags *p*, corresponding to 7, 14 and 21 days.

#### 2.4.4 LSTM

We selected a stateful long short-term memory (LSTM) network [42] as a simple yet flexible deep learning model to capture temporal dependencies in the time-series data. In this configuration, hidden states are carried over between consecutive input sequences rather than reset after each batch. The model comprised three LSTM layers with 50 units each, each followed by a dropout layer (rate 0.2) to reduce overfitting. The output layer was a dense layer with a single unit, wrapped in a *TimeDistributed* layer to produce one output for each input time step.

### 2.5 Performance evaluation and model baselines

Model performance was evaluated using RMSE and Symmetric Mean Absolute Percentage Error (SMAPE), a scale-independent variant of Mean Absolute Percentage Error (MAPE). While MAPE measures the mean relative difference between actual and predicted values, it disproportionately weights errors during periods of low incidence, since it divides by the actual value. SMAPE mitigates this imbalance by normalizing errors by the mean of the absolute actual and predicted values.

As a baseline, we repeated the procedure using only the time variable as a predictor and compared its performance to the LASSO and MLP models. For time-aware models (VAR and LSTM), the current disease incidence served as the sole predictor, with temporal information captured implicitly through the sequential ordering of the data.

### 2.6 Differential privacy

We employed Differential Privacy (DP) [48] to ensure that individual-level data contributions could not be inferred from the model outputs. DP provides a formal guarantee that the inclusion or exclusion of a single data point has a limited effect on the analysis, achieved by injecting calibrated noise during training. The strength of the privacy guarantee is controlled by the parameters *ε* (privacy loss) and *δ* (failure probability), with smaller values corresponding to stronger privacy.

DP can be applied at the data level or during model training. In this study, we focus on model-level privacy using Differentially Private Stochastic Gradient Descent (D-SGD) [49], a widely adopted method for training ML models with DP guarantees. A key contribution is our implementation of DP-SGD in R, making state-of-the-art privacy-preserving training accessible to researchers beyond the core machine-learning community.

In addition, we quantify the resulting privacy guarantees using Rényi Differential Privacy [50] (RDP), which offers a tighter and more flexible way to track cumulative privacy loss under composition compared with standard DP analysis. More details on the privacy implementation can be found in the Supplementary data, Section S1.

### 2.7 Explainability

The development of explainable ML methods is crucial for identifying the input features that most strongly influence model predictions, addressing concerns associated with black-box approaches, and forming a central aim of this study.

For LASSO and VAR, feature importance was derived directly from estimated coefficients obtained from models trained on the first 90% of time-ordered data.

For deep learning models, we applied two post-hoc, model-agnostic explainability methods, Local Interpretable Model-agnostic Explanations (LIME) [51] and SHapley Additive exPlanations (SHAP) [52], to ensure robustness. Both provide local explanations by attributing importance to individual predictions: LIME fits a simple surrogate model locally using perturbed inputs, while SHAP uses game-theoretic Shapley values to quantify each feature’s contribution in an additive, theoretically grounded manner. LIME and SHAP were computed using the R package *iml* [53] for the MLP, and the Python packages *lime* and *shap* [54] for the LSTM.

#### 2.7.1 Clustering and aggregation for SHAP/LIME

To facilitate SHAP and LIME analyses, the standardized dataset was clustered into five groups based on incidence using k-means with the elbow method. Explanations were computed for the two clusters with the lowest and highest incidences, while the intermediate clusters were omitted to focus on clear contrasts between low-and high-incidence regimes. In low-incidence clusters, influential features that are positively associated with incidence typically have negative contributions, reflecting feature-specific effects on individual predictions as captured by LIME/SHAP, i.e., local reductions in predicted incidence relative to the baseline. Conversely, in high-incidence clusters, such features tend to contribute positively, increasing the predicted incidence for individual samples.

Feature importance was averaged across all samples within each cluster. For the LSTM, attributions were computed per feature and per input–output time step. To align with the forecasting horizon (day 7), explanations were generated using a model trained on data ending eight days earlier. Importance values were then averaged across the seven input steps to obtain a single feature importance profile per cluster.

#### 2.7.2 Interpreting SHAP values

In the SHAP-based tables, feature effects are reported relative to the model’s baseline prediction, defined as the expected prediction over all samples. For each cluster, feature values are approximated by the cluster mean and compared to the overall feature mean to indicate deviation from the background distribution, shown in the tables with an arrow next to the mean SHAP value (upward if above the overall mean, downward if below).

A SHAP value quantifies whether a feature locally increases or decreases predicted incidence relative to the baseline. For roughly monotonic relationships, the implied direction of association depends on both the SHAP sign and whether the cluster-level feature value is above or below its overall mean. For example, a negative SHAP value for a below-average feature indicates a positive association, as lower feature values are associated with lower predicted incidence.

This interpretation follows from the definition of SHAP values, which measure marginal feature contributions averaged over all possible coalitions relative to the empirical background distribution. Consequently, SHAP attributions are inherently relative and should not be interpreted as absolute effect sizes.

#### 2.7.3 Interpreting LIME values

LIME explanations represent local feature effects for individual samples, computed as the product of the feature value and the coefficient of the locally fitted surrogate model. For features with a negative association and strictly positive scale (e.g., body mass or counts), LIME effects remain negative, varying only in magnitude, so their sign does not indicate whether the feature is above or below its population mean, unlike SHAP values, which are defined relative to a baseline and can change sign based on deviation from the reference.

Consequently, comparing LIME effects to population means is not meaningful when raw feature scales are used. However, if features are standardized to zero mean and unit variance prior to fitting the LIME surrogate model, as described in Section 2.3, feature values directly encode deviations from the mean. In this setting, the sign of the LIME effect reflects whether a feature value is above or below average, and under locally approximately monotonic relationships this yields an interpretation qualitatively comparable to SHAP. This equivalence, however, relies on the assumption that the local linear approximation captures the underlying monotonic relationship.

#### 2.7.4 Assessing the impact of differential privacy

Finally, we assessed the impact of differential privacy (DP) on feature importance in COVID-19 incidence prediction. A simplified procedure was used for MLP models to evaluate the stability of explainability values across different privacy levels, rather than to achieve detailed interpretability. For each privacy budget, 10-fold cross-validation was performed: in each fold, the MLP was trained on the training subset, and absolute explainability values were computed on the corresponding validation subset. The results were then averaged across folds to produce a stable aggregate measure of feature importance for each privacy budget.

## 3 Results

### 3.1 Predictive performance

The seven-day prediction results on the full dataset are summarized in Table 1 (RMSE) and Table 2 (SMAPE). For each model, results are presented separately for training and testing, with the MuSPAD-based model on the left and the baseline on the right. Fig. 1 shows predicted and observed COVID-19 incidence across models.

**Table 1:** RMSE for seven-day predictions using a full-data split into training and testing.

| Time-agnostic methods |  |  |  |  |
| --- | --- | --- | --- | --- |
| Method | Training MuSPAD | Test MuSPAD | Training baseline | Test baseline |
| LASSO | 33.09 | 9.28 | 70.73 | 61.70 |
| MLP | 30.52 | 15.13 | 75.51 | 33.65 |
| Time-aware methods |  |  |  |  |
| Method | Training MuSPAD | Test MuSPAD | Training baseline | Test baseline |
| LSTM | 31.34 | 4.36 | 68.84 | 3.94 |
| VAR $p=7$ | 34.59 | 36.16 | 38.45 | 30.49 |
| VAR $p=14$ | 36.93 | 15.54 | 38.80 | 30.41 |
| VAR $p=21$ | 36.14 | 11.00 | 39.42 | 27.18 |

The left column lists the models evaluated, while the remaining columns show the corresponding RMSE values. “MuSPAD” indicates that the model includes the MuSPAD variables in its predictions, while “baseline” refers to the respective baseline models described in Section 2.5.

**Table 2:** SMAPE for seven-day predictions using a full-data split into training and testing.

| Time-agnostic methods |  |  |  |  |
| --- | --- | --- | --- | --- |
| Method | Training MuSPAD | Test MuSPAD | Training baseline | Test baseline |
| LASSO | 0.39 | 1.07 | 0.91 | 1.46 |
| MLP | 0.48 | 0.76 | 1.06 | 1.20 |
| Time-aware methods |  |  |  |  |
| Method | Training MuSPAD | Test MuSPAD | Training baseline | Test baseline |
| LSTM | 0.54 | 0.37 | 0.88 | 0.29 |
| VAR $p=7$ | 0.62 | 1.26 | 0.51 | 1.13 |
| VAR $p=14$ | 0.66 | 0.84 | 0.49 | 1.13 |
| VAR $p=21$ | 0.64 | 0.68 | 0.48 | 1.07 |

The remaining notation is as in Table 1.

**Figure 1:**
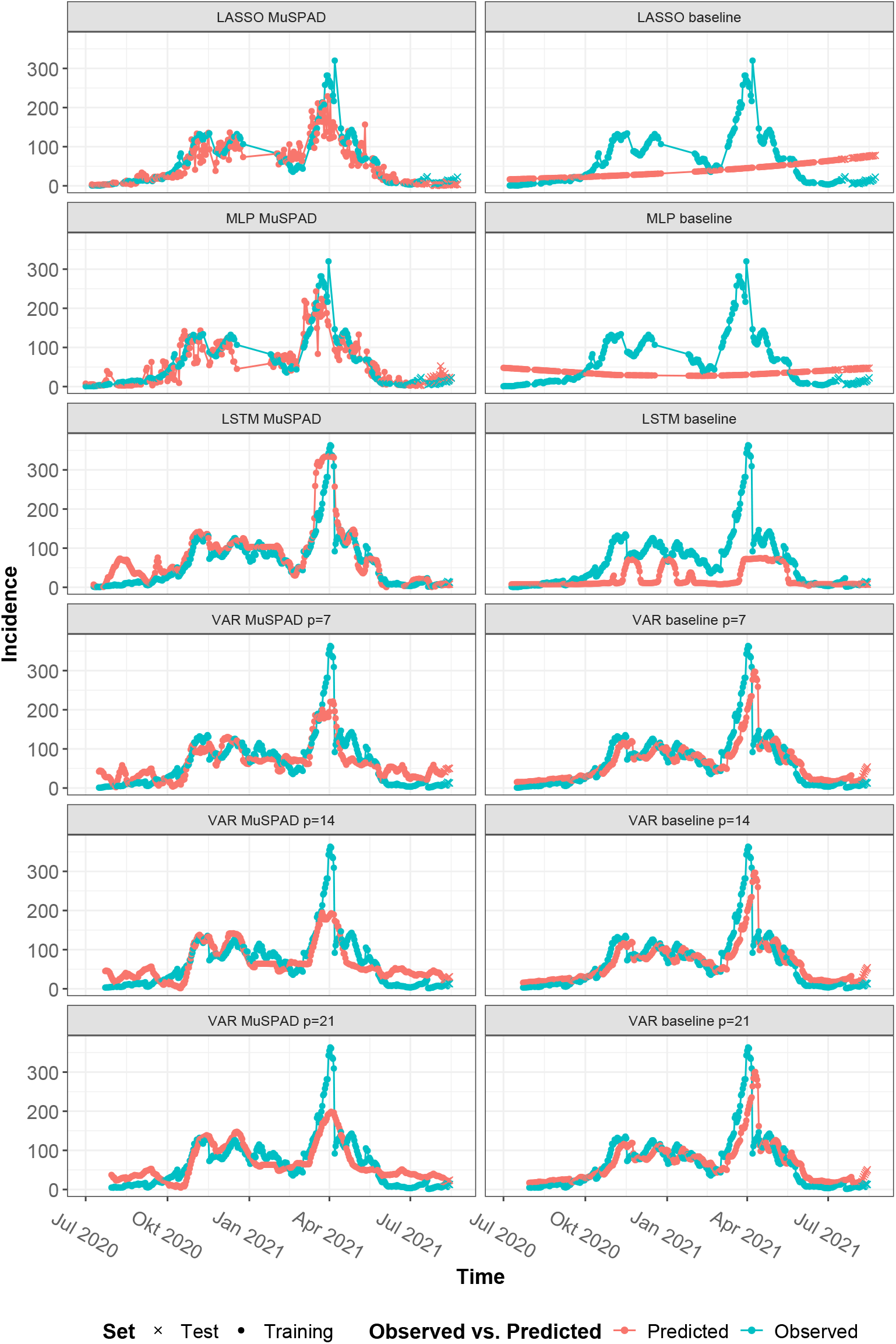
Observed vs. Predicted COVID-19 Incidence Across Models (Full Data). Comparison of all observed (blue) versus models’ predicted (red) COVID-19 incidences for training (points) and testing (crosses) in seven-day predictions for the full dataset, with models including MuSPAD variables (left) versus baseline models described in Section 2.5 (right). Model types are indicated in the respective plot title; for VAR model *p* and *h* correspond to lag length and prediction horizon respectively. Evaluation metrics are reported in Tables 1 and 2.

Incorporating MuSPAD features substantially improved performance for the non–time-aware models (LASSO and MLP), reducing both RMSE and SMAPE relative to the baselines in training and testing (e.g., LASSO RMSE: 33 vs. 71 in training and 9 vs. 62 in testing). Among time-aware models, LSTM with MuSPAD features achieved strong test performance (RMSE 4.36; SMAPE 0.37), comparable to or slightly better than the baseline, while VAR models exhibited lag-dependent effects: the MuSPAD variant with *p* = 7 lag underperformed on the test set, whereas *p* = 14 and *p* = 21 outperformed their baselines in both RMSE and SMAPE. Overall, the LSTM achieved the lowest test error, while LASSO and MLP showed lower training errors, reflecting differences in model complexity and regularization.

Fig. 1 shows that the time-agnostic MuSPAD models captured the overall trend well but display oscillations between time points due to the lack of temporal context. On the test set, LASSO achieved a slightly better fit than MLP, likely because its simpler structure and stronger regular-ization allow it to closely track short-term fluctuations in this limited dataset without overfitting. Baseline models using only time as a predictor reduced to a simple, slightly curved trend line.

From around November 2020 onward, the MuSPAD LSTM model (Fig. 1, third row, left) yielded a smoother fit. It accurately captured the large April 2021 wave, being the only model to approach the observed peak. In the initial observation period (before October 2020), however, it predicted spurious waves not reflected in the actual data.

The MuSPAD VAR models (Fig. 1, last four rows, left) captured many of the smaller waves but tended to produce flatter predictions than observed. Increasing the time lag *p* smoothed the fitted curve and improved test performance. In contrast, the VAR baseline models (last four rows, right), which used seven days of input to predict the following week, largely replicated the pattern from about seven days earlier. While the initial fit appeared reasonable, large deviations occurred when high incidences followed a decline. After August 2021, the model often predicted rises that did not materialize. Similarly, the baseline LSTM forecasted incidence waves with a delay, substantially underestimated peak values, and shown low variability, yielding deceptively low error metrics by effectively predicting near-constant incidence.

When trained on three-quarters of the data and tested on the subsequent seven time points (Table 3), the MuSPAD LSTM model exhibited a clearer advantage over the baseline, achieving both better training and test performance compared with the full-data analysis. For VAR models, the earlier benefit of MuSPAD at *p ≥* 14 disappeared, even though the baseline shown much higher training RMSE than the MuSPAD variants (Fig. 2). The baseline models failed to capture underlying trends in the training data but still achieved reasonable test performance by effectively predicting a near-constant incidence equal to the global average.

**Table 3:** RMSE for seven-day predictions using a three quarters of the data split into training and testing.

| Time-aware methods |  |  |  |  |
| --- | --- | --- | --- | --- |
| Method | Training MuSPAD | Test MuSPAD | Training baseline | Test baseline |
| LSTM | 22.75 | 26.88 | 129.10 | 39.52 |
| VAR p=7 | 33.80 | 36.58 | 53.04 | 2.51 |
| VAR p=14 | 31.19 | 29.47 | 53.31 | 2.84 |
| VAR p=21 | 34.59 | 36.16 | 67.21 | 14.89 |

The remaining notation is as in Table 1.

**Figure 2:**
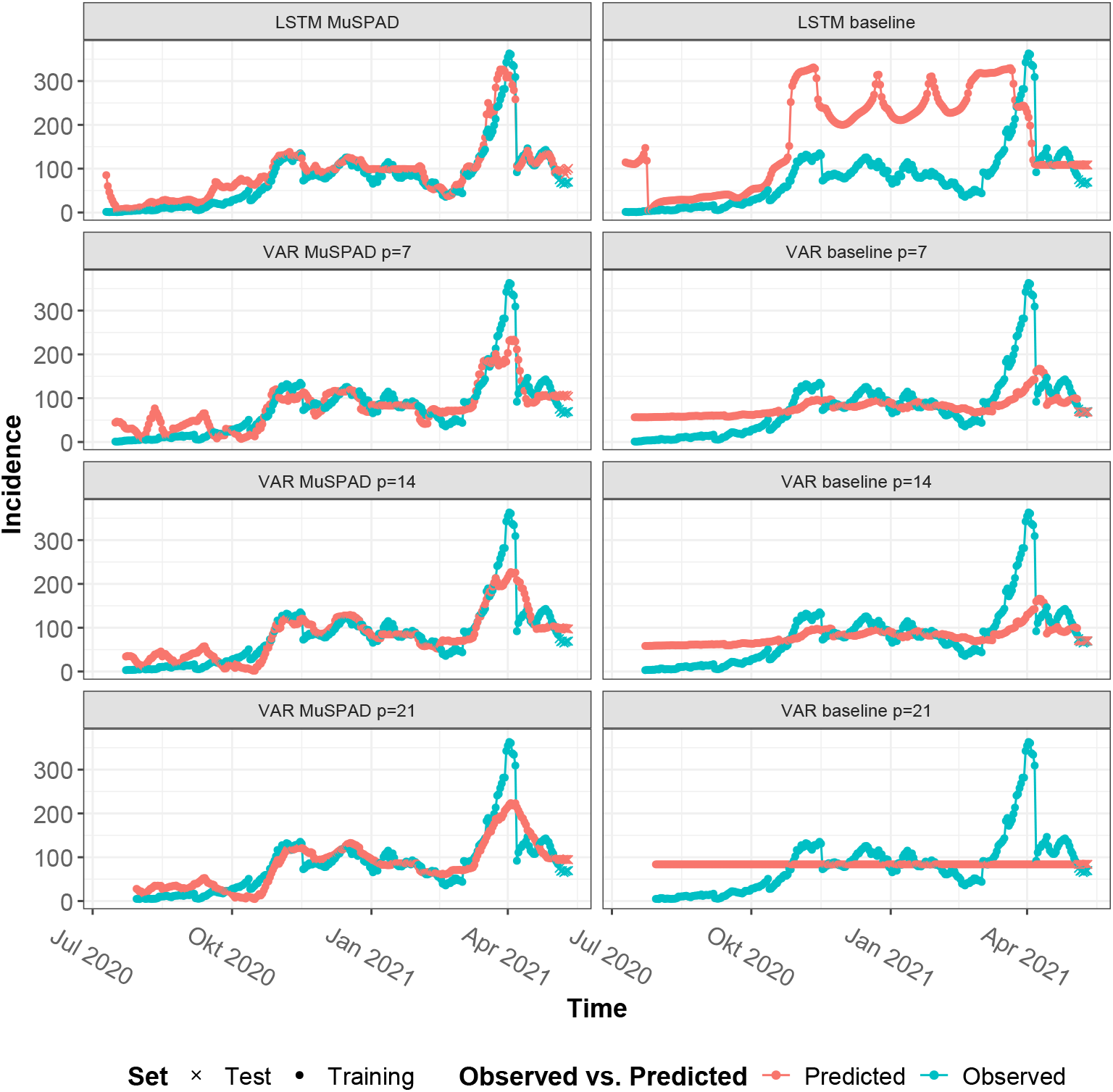
Observed vs. Predicted COVID-19 Incidence Across Models (First 75% of the Data). Comparison of all observed (blue) versus models’ predicted (red) COVID-19 incidences for training (points) and testing (crosses) in seven-day predictions, with MuSPAD data (left) versus baseline models (right). The first three-quarters of the time points were used as training data. Corresponding evaluation metrics are reported in Table 3. The remaining notation is as in Fig. 1.

### 3.2 Explainability

Feature importance is summarized in Tables 4–7,S1 and S2, reporting the top 10 predictors by absolute importance across models. Regression-based models report coefficients (Tables 5 and 7), while deep learning models are explained via SHAP and LIME for MLP and LSTM separately (Tables 4, 6, S1 and S2), with results shown for high-and low-incidence clusters. Across models and explainability methods, several variables were consistently ranked among the most influential predictors of COVID-19 incidence, demonstrating relevance across approaches.

**Table 4:** Explainability table of Shapley values for the 10 most important features (ranked by absolute value) in low-and high-incidence clusters for the MLP model.

| MLP low Feature Title | Shapley low | MLP high Feature Title | Shapley high |
| --- | --- | --- | --- |
| No mask at restaurant | -2.09 (↓) | No mask at restaurant | 9.24 (↑) |
| Antibodies reactive to SARS-CoV-2 Spike S1 | -1.64 (↑) | Serology status Infected | 5.77 (↑) |
| Received second vaccination | -1.29 (↑) | Tested PCR positive | 5.74 (↑) |
| Serostatus vaccinated second | -1.13 (↑) | Covid19 test at least once positive since 02/2020 | 5.47 (↑) |
| Fully vaccinated | -1.11 (↑) | Someone in household PCR tested positive | 5.40 (↑) |
| Antibodies reactive to SARS-CoV-2 Spike protein | -1.00 (↑) | Someone in household tested positive since 02/2020 | 5.31 (↑) |
| Missing information on PCR testing | -1.00 (↓) | Contact with confirmed COVID 19 patient | 4.99 (↑) |
| Missing information on maintenance of social distance | -0.95 (↓) | Natural immunity | 4.88 (↑) |
| No positive PCR test | -0.93 (↓) | Serostatus natural immunity | 4.76 (↑) |
| Employment changes during pandemic | -0.90 (↑) | PCR tested, once positive | 4.49 (↑) |

“MLP low Feature Title” and “Shapley low” contain information on low incidence cluster; “MLP high Feature Title” and “Shapley high” contain information on high incidence cluster. (↓) indicates that mean of feature values in cluster (feature cluster mean) is below overall mean of feature values (feature overall mean); (↑) indicates that feature cluster mean is above feature overall mean.

**Table 5:** Explainability table of 10 highest coefficients (ranked by absolute value) in LASSO model.

| LASSO Feature Title | LASSO Coefficient |
| --- | --- |
| Number of days since 01st Jan 2020 | 1.15 |
| Antibodies reactive to SARS-CoV-2 Receptor-binding domain | -0.75 |
| Employment changes during pandemic, | -0.46 |
| Antibodies reactive to SARS-CoV-2 Spike protein | -0.36 |
| Mask at sports | 0.25 |
| No employment changes during pandemic | 0.19 |
| Count of all symptoms since 02/2020 | 0.17 |
| Mask in public places | 0.15 |
| Antibodies reactive to SARS-CoV-2 Spike S1 | -0.15 |
| Fully maintenance of social distance | -0.13 |

**Table 6:** Explainability table of Shapley values for the 10 most important features (ranked by absolute value) in low-and high-incidence clusters for the LSTM model.

| LSTM low Feature Title | Shapley low | LSTM high Feature Title | Shapley high |
| --- | --- | --- | --- |
| seven-day-Incidence of COVID-19 per 100,000 based on RKI-Notifications | -0.95 (↓) | seven-day-Incidence of COVID-19 per 100,000 based on RKI-Notifications | 3.19 (↑) |
| Missing information on quarantine status | -0.28 (↓) | No mask at restaurant | 2.14 (↑) |
| Missing information on Maintenance of social distance | -0.27 (↓) | Quarantine status: mandatory / voluntary quarantine | -2.02 (↑) |
| No mask at restaurant | -0.26 (↓) | Hand washing 60 sec. recommended time | 1.78 (↑) |
| Missing information on PCR testing in the household | -0.24 (↓) | Natural immunity | -1.69 (↑) |
| Hand washing 60 sec. recommended time | -0.22 (↓) | Covid19 test since 02/2020 positive at least once | -1.69 (↑) |
| Missing information on PCR testing | -0.21 (↓) | No maintenance of social distance | 1.61 (↑) |
| Mask go for a walk | -0.20 (↓) | Missing information on PCR testing in the household | 1.37 (↑) |
| Mask at other occasions | -0.19 (↓) | Mask to go for a walk | 1.21 (↑) |
| Euroimmun result negative | -0.18 (↓) | Employment changes during pandemic: parental or other leave | 1.19 (↑) |

Notation as in Table 4.

**Table 7:** Explainability table of 10 highest coefficients (ranked by absolute value) in VAR model.

| VAR Feature Title | VAR Coefficient |
| --- | --- |
| seven-day-Incidence of COVID-19 per 100,000 based on RKI-Notifications | 0.0937 |
| No mask at restaurant | 0.0915 |
| Natural immunity | 0.0906 |
| Serostatus natural immunity | 0.0902 |
| Contact with confirmed COVID 19 patient | 0.0789 |
| No mask public transport | 0.0758 |
| Mask public places | 0.0755 |
| Serology status Infected | 0.0744 |
| Tested PCR positive | 0.0742 |
| PCR tested, once positive | 0.0742 |

To further structure the analysis, features were grouped into six categories: Contacts and Non-Pharmaceutical Interventions (NPIs), Employment, Immunity, Masks, Testing, and Vaccination (as shown in Figs. 3 and 4). Considering the top 50 absolute explainability values, testing-and employment-related features consistently emerge among the highest contributors across all models.

**Figure 3:**
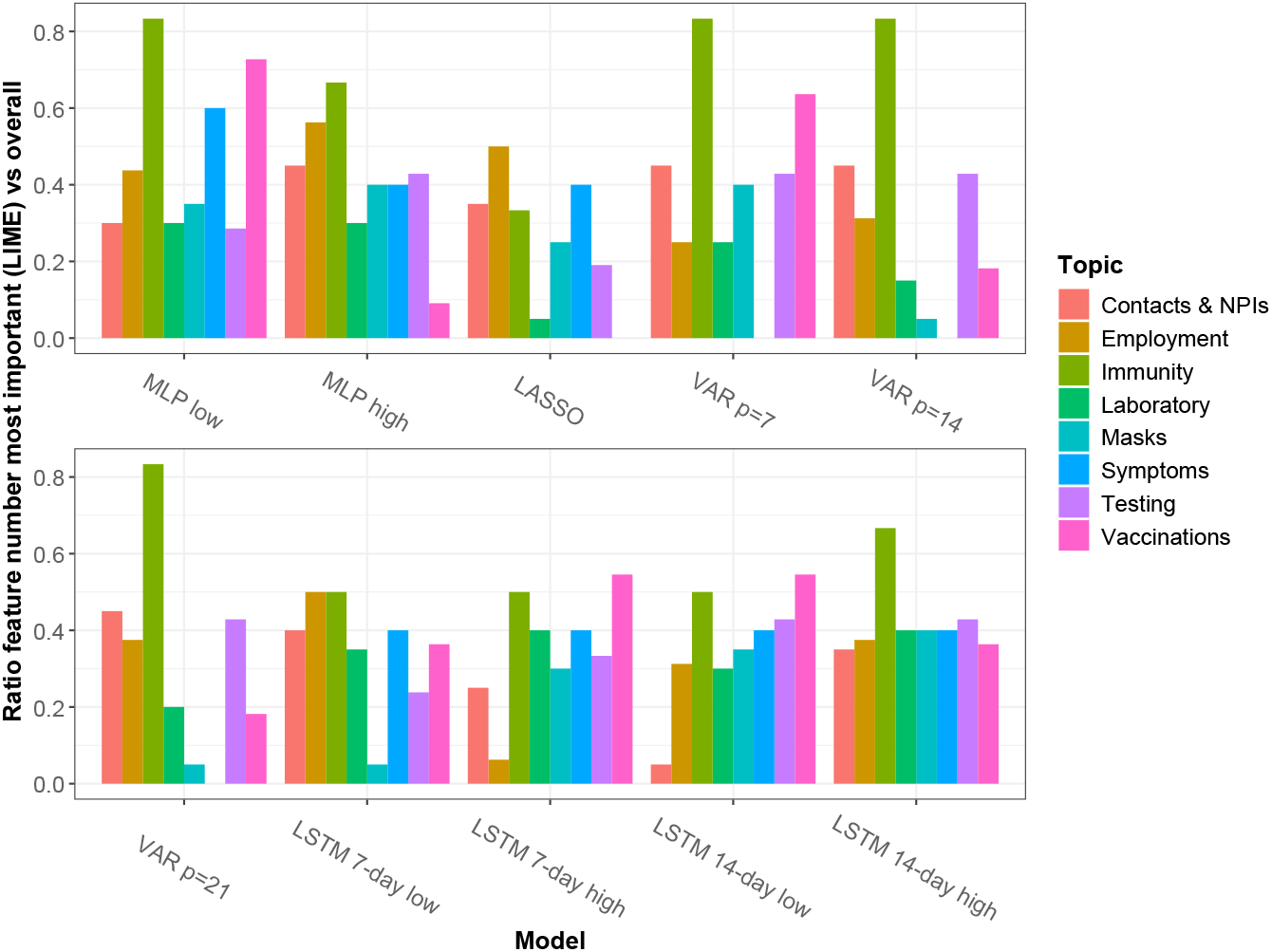
Distribution of top-50 features by topic and model (LIME). Bar plots show the proportion of features among the 50 most important features (ranked by absolute value) relative to the total number of features, stratified by topic and model, as identified by LIME explainability.

**Figure 4:**
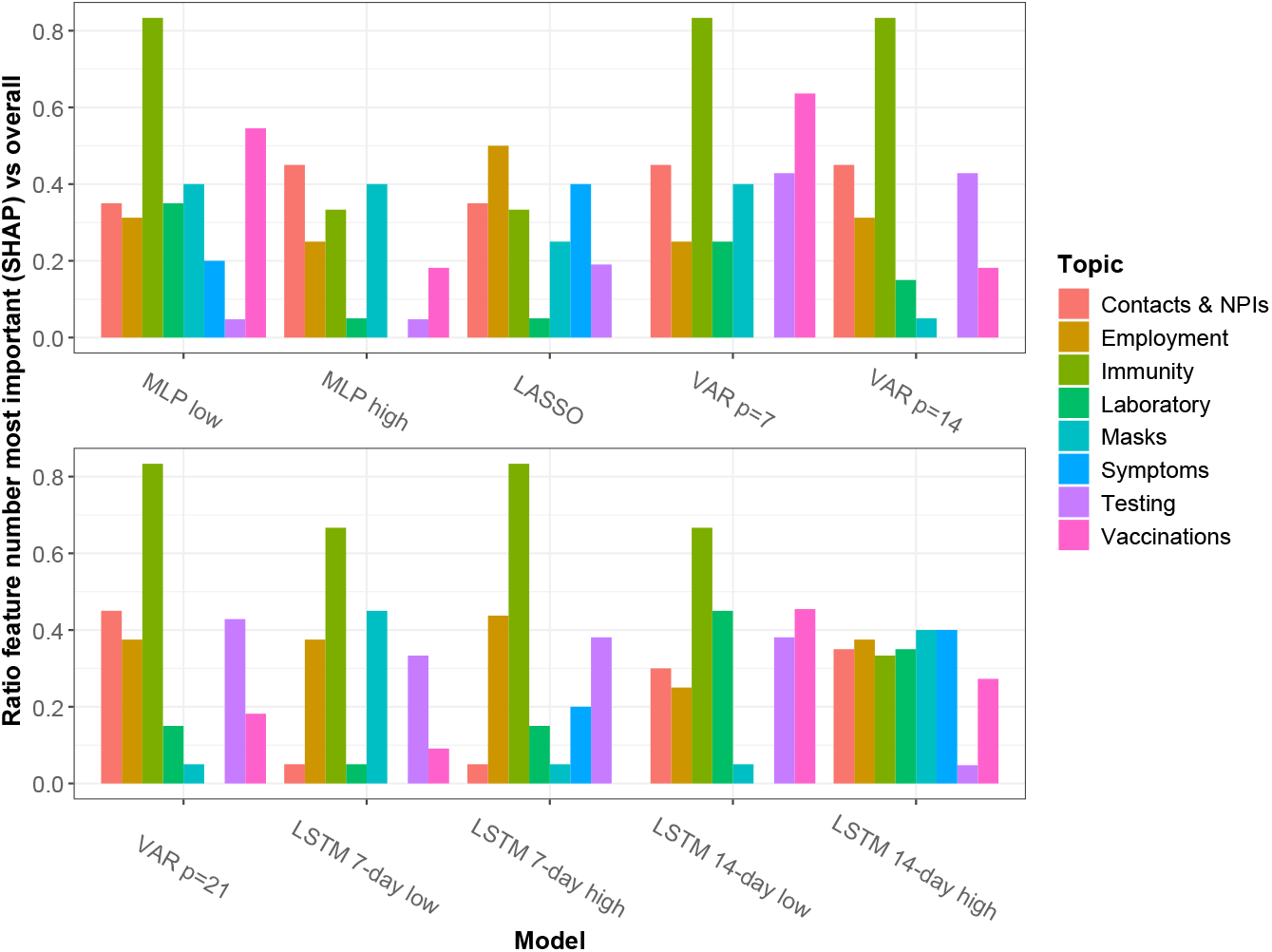
Distribution of top-50 features by topic and model (SHAP). Bar plots show the proportion of features among the 50 most important features (ranked by absolute value) relative to the total number of features, stratified by topic and model, as identified by SHAP explainability.

#### 3.2.1 Key explainability patterns

Most notably, “No mask at restaurant” was a key feature across all model classes. In the MLP, it shown a strong negative contribution in the low-incidence cluster (SHAP: -2.09; LIME: -4.39; *↓* cluster mean, Tables 4, S1) and a strong positive contribution in the high-incidence cluster (SHAP: 9.24; LIME: 20.32; *↑* cluster mean). A similar pattern appeared in the LSTM, with negative SHAP and LIME values in the low-incidence cluster (−0.26 and -1.13 respectively; *↓*, cluster mean, Tables 6, S2) and positive SHAP values in the high-incidence cluster (2.14; *↑* cluster mean). In the VAR model, it had one of the largest positive coefficients (0.0915). Together, these results indicated that not wearing masks at restaurants was consistently associated with higher predicted incidence across both local and global models.

Other mask-related variables, including “Wearing mask in public places,” “Mask to go for a walk,” and “Mask at home,” also shown consistent associations: higher reported mask use corresponded to higher predicted incidence. For example, in the MLP LIME low-incidence cluster, “Wearing mask in public places” had a strong negative contribution (−11.85, *↓*, Table S1), indicating that below-average use predicted lower incidence, while above-average use predicted higher incidence. In high-incidence clusters, these variables contributed positively (*↑*). In contrast to “No mask at restaurant,” this pattern likely reflects behavioral responses to increasing incidence rather than a direct risk signal. This difference may reflect that “No mask at restaurant” captures a specific high-risk exposure setting, whereas other mask-related variables primarily reflect behavioral responses to changing incidence.

Infection history and testing variables were consistently ranked among top predictors. In the MLP SHAP high-incidence cluster, “Tested PCR positive,” “PCR tested, once positive,” and “Serology status Infected” had SHAP values 4.49–5.77 (Table 4, *↑*), while in VAR their coefficients were 0.074–0.078 (Table 7), indicating that higher prevalence of prior infections predicted higher incidence.

Missing PCR information was also influential, particularly in low-incidence clusters (MLP SHAP: -1.00 *↓*; LSTM SHAP: -0.21 to -0.24 *↓*, Tables 4, 6), reflecting that incomplete reporting generally associated with higher predicted incidence. Overall, infection/testing history and reporting patterns were meaningful predictors across both temporal and non-temporal models.

Employment-related variables were important predictors across models, with contributions varying by cluster and model type. In the MLP, “Employment changes during pandemic” shown a negative LIME value in the low-incidence cluster (−0.90, *↓*, Table S1) and a positive value in the high-incidence cluster (10.43, *↑*, Table S1), both reflecting positive associations with predicted incidence. In the LSTM, low-incidence clusters shown positive associations for “Employment changes” (−0.98) and “No employment changes” (−1.26, *↓*, Table S2), while in high-incidence clusters “Employment changes” (3.06, *↓*, Table S2) was negatively associated, and “Parental/other leave” (SHAP 1.19, *↑*) and “No employment changes” (2.97, *↑*) are positively associated.

In LASSO, coefficients for “Employment changes” (−0.46) and “No employment changes” (0.19) mirror the high-incidence LSTM patterns. The largest LASSO coefficient, however, was for lagged incidence, which captured historical effects (e.g., vaccination) and may have reduced the apparent importance of other predictors, though the model still outperformed a baseline using only past incidence.

Immunity-related variables demonstrated model-specific pattern, including e.g. “Antibodies reactive to SARS-CoV-2 Spike protein”, “Spike S1”, and “Receptor-binding domain”. In the non-temporal models, these features generally contributed negatively to predicted incidence. For instance, in the MLP SHAP low-incidence cluster, “Antibodies reactive to SARS-CoV-2 Spike S1” had a SHAP value of -1.64 with an *↑* arrow (Table 4), indicating that although the cluster mean was higher than the overall mean, the feature locally decreased predicted incidence. Similarly, in the MLP LIME low-incidence cluster, “Antibodies reactive to SARS-CoV-2 Receptor-binding domain” shown a negative LIME value of -5.58 with an *↑* arrow (Table S1), and in LASSO, the coefficients for “Spike S1” (−0.15) and “Receptor-binding domain” (−0.75, Table 5) also reflected a globally negative contribution.

In contrast, temporally aware models shown more complex behavior for immunity-related variables. Neither the VAR nor the LSTM ranked antibody-related variables among the top 10 predictors, and several antibody variables appearing in the LSTM top 50 (Supplementary Data) were positively associated with incidence. Similarly, in the VAR model, “Natural immunity” and “Serostatus natural immunity” had positive coefficients (0.0906 and 0.0902, Table 7), suggesting that higher-than-average immunity levels were associated with increased predicted incidence, likely reflecting temporal correlations with past incidence trends rather than a direct protective effect.

These patterns shown that temporally aware models integrated past incidence and evolving immunity, making the local effect of immunity features dependent on both cluster value and temporal context, whereas non-temporal models treat immunity mainly as a cross-sectional indicator of lower contemporaneous incidence.

### 3.3 Differential privacy

Figs. 5 and 6 show how LIME-and SHAP-derived feature importance values vary across privacy budgets for six representative features.

**Figure 5:**
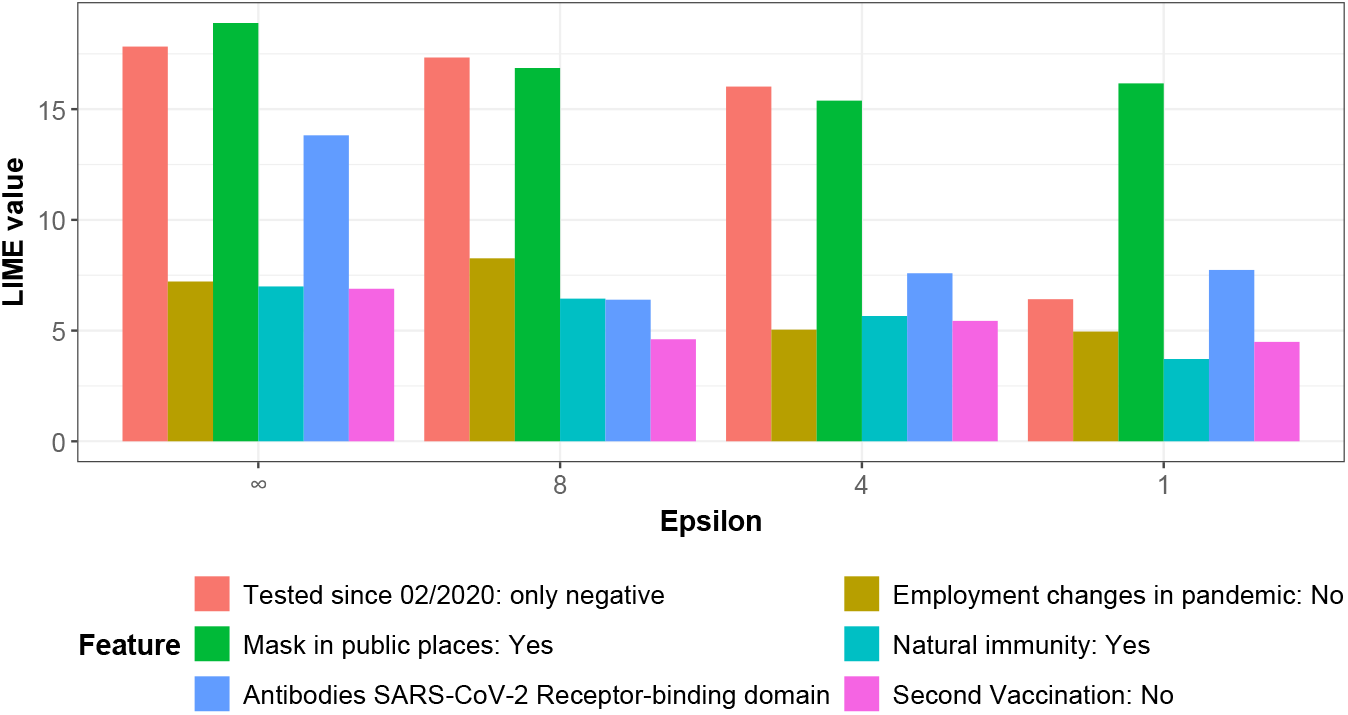
LIME feature importance across privacy budgets (MLP). Bar plots show absolute LIME values (vertical axis) for selected features across different privacy budgets (*ε*), compared with the non-private model (horizontal axis) in the MLP.

**Figure 6:**
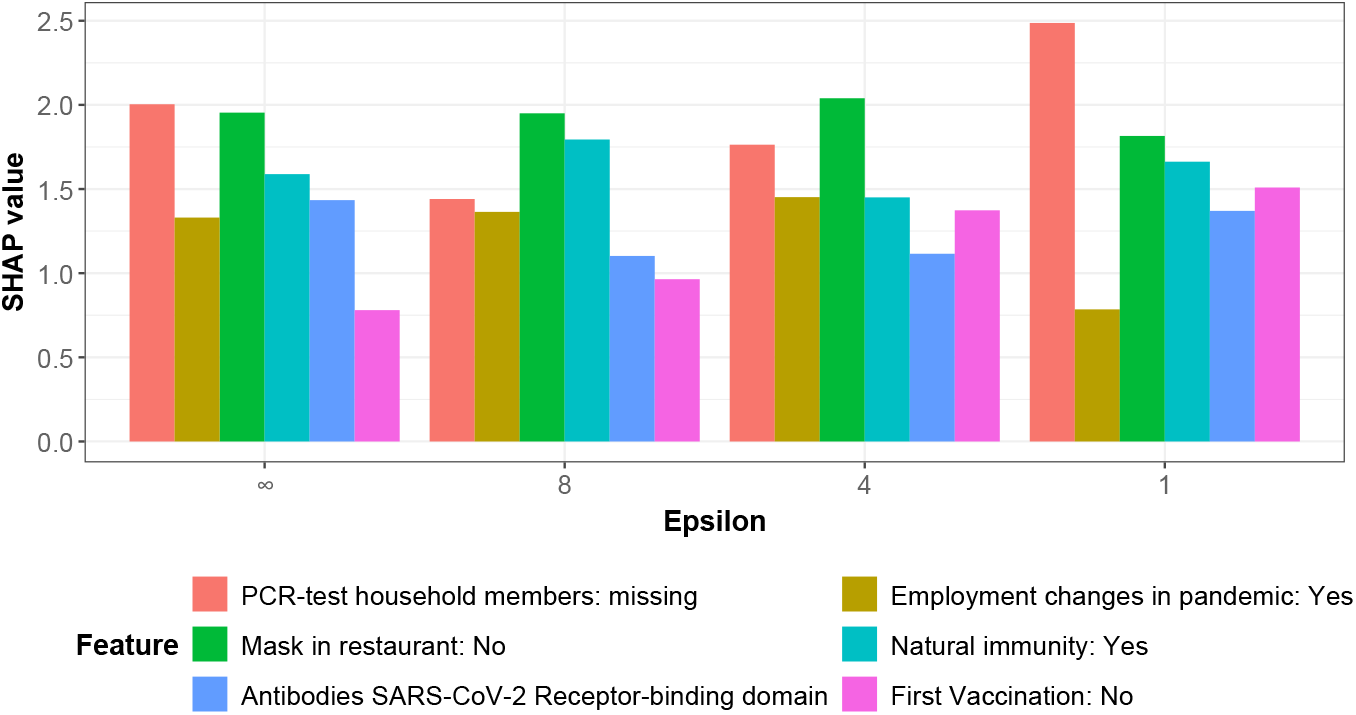
SHAP feature importance across privacy budgets (MLP). Bar plots show absolute SHAP values (vertical axis) for selected features across different privacy budgets (*ε*), compared with the non-private model (horizontal axis) in the MLP.

Table 8 illustrates the privacy–utility trade-off induced by differential privacy. As the privacy budget *ε* decreased (corresponding to stronger privacy), training RMSE increased monotonically, reflecting the growing impact of DP noise on model fitting. Validation RMSE shown a non-monotonic pattern: moderate privacy budgets (*ε* = 8 and *ε* = 4) yielded validation errors comparable to or lower than the non-private baseline, suggesting a regularization effect that can improve generalization. However, at stronger privacy budgets (*ε* = 1), both training and validation performance deteriorated substantially, indicating that excessive noise overwhelms the predictive signal.

**Table 8:** RMSE for seven-day incidence predictions obtained with the MLP model under varying levels of differential privacy.

| $\epsilon$ | training RMSE | validation RMSE |
| --- | --- | --- |
| ∞ | 977.57 | 1337.32 |
| 8 | 1031.29 | 1224.54 |
| 4 | 1231.46 | 1257.08 |
| 1 | 1684.31 | 1970.82 |

Results are reported separately for training and validation sets. Differential privacy strength is controlled by the privacy budget parameter $\epsilon$ , with smaller values corresponding to stronger privacy guarantees. The case $\epsilon = \infty$ denotes the non-private baseline without differential privacy.

For LIME, smaller privacy budgets were associated with a decline in feature importance, whereas for SHAP, although some oscillations were observed across privacy budgets, the overall magnitude of feature importance remains relatively stable.

Conceptually, this difference can be explained by the distinct mechanisms underlying LIME and SHAP. LIME approximates the model’s local behavior by fitting a simple surrogate model to perturbed samples around each instance. When DP is applied, the injected noise disrupts these local perturbations, reducing the fidelity of the surrogate model and leading to a decline in measured feature importance at smaller privacy budgets. In contrast, SHAP computes feature contributions based on the average marginal effect of each feature across many coalitions of inputs. This aggregation process smooths out the impact of DP-induced noise, resulting in oscillations across privacy budgets but overall stable importance values. Thus, LIME’s local sensitivity makes it more affected by privacy noise, whereas SHAP’s global averaging confers greater robustness.

## 4 Discussion

In this study, we show that aggregated individual-level information from the MuSPAD seroprevalence cohort can predict local seven-day COVID-19 incidence with reasonable accuracy when appropriate model classes and configurations are used. Beyond predictive performance, our results provide insight into how time-aware and time-agnostic models leverage epidemiological, behavioral, and available socioeconomic signals, and how these are reflected in explainability outputs under differential privacy, highlighting the value of combining heterogeneous data sources in digital health settings.

Across most models and explainability methods, variables related to past infections, employment, and testing activity emerged as consistent predictors of incidence. Features capturing PCR positivity, prior infection, and serological infection status consistently showed positive contributions in high-incidence clusters and positive global coefficients in the VAR model. This pattern likely reflects the strong coupling between recent transmission dynamics, testing intensity, and observed incidence. At the same time, variables encoding missing PCR information were frequently influential, particularly in low-incidence clusters, where lower-than-average missingness tended to reduce predicted incidence. This suggests that reporting behavior itself carries predictive information, likely reflecting differential engagement with testing and surveillance during different epidemic phases.

Mask-related variables highlight the interaction between behavioral responses and model structure. When stratifying observations by incidence level, the feature “No mask at restaurant” shown a consistent and interpretable pattern: lower values were associated with reduced predicted incidence in low-incidence settings, whereas higher values increased predictions in high-incidence settings, consistent with a direct risk signal. This pattern is strongest in non-temporal models and is partially attenuated in temporal models through the inclusion of lagged incidence, indicating partial overlap between behavioral signals and temporal dynamics.

Employment-related variables are consistently relevant across models but exhibit clear differences between non-temporal and time-aware approaches. In non-temporal models, particularly MLP and LASSO, above-average employment disruptions are associated with higher predicted incidence, especially in high-incidence clusters, likely reflecting reactive responses to NPIs such as workplace closures rather than direct exposure effects. In contrast, temporal models, most notably the LSTM, capture more context-dependent and sometimes opposing local effects, indicating how different forms of employment change (e.g., leave, job loss, or stability) interact with past incidence and policy dynamics. Overall, employment variables function both as markers of NPI-driven behavior and as proxies for broader socioeconomic disruption during epidemic waves.

Differences between model classes are most evident for immunity-related variables. In nontemporal models (LASSO and MLP), antibody-related features consistently show negative contributions to predicted incidence, aligning with their interpretation as contemporaneous indicators associated with lower incidence. In contrast, temporally aware models assign limited and qualitatively different importance to these variables. Antibody measures rarely rank among the top predictors in VAR or LSTM and, when they do, their associations are often positive. In particular, VAR assigns positive coefficients to “Natural immunity” and “Serostatus natural immunity,” indi cating that higher immunity levels coincide with higher predicted incidence. This contrast suggests that temporal models primarily capture immunity variables through their correlation with past incidence dynamics rather than as direct modifiers of current risk, highlighting the role of temporal confounding in interpreting such associations.

The feature group analysis further confirmed the prominence of testing and employment-related features among the top contributors across models. This finding emphasizes that behavioral engagement with surveillance systems and socioeconomic conditions are central to explaining observed variation in incidence, and that such signals are typically absent from routine surveillance systems, consistent with prior epidemic modeling studies [55].

Finally, the differential privacy results highlight clear trade-offs between privacy, predictive performance, and interpretability. As the privacy budget decreases, training error increases monotonically, reflecting reduced model capacity under stronger privacy constraints, while validation error exhibits a non-monotonic pattern consistent with a regularization effect at moderate privacy budgets and substantial degradation under stronger privacy budgets (*ε* = 1). In terms of interpretability, SHAP attributions remain comparatively stable across privacy levels, whereas LIME becomes increasingly unstable as privacy noise increases. This suggests that global, aggregation-based explanations such as SHAP are better suited for differentially private settings than local, perturbation-based methods in privacy-sensitive digital health applications.

Several limitations should be considered. First, the MuSPAD data are based on repeated cross-sectional sampling rather than longitudinal follow-up of the same individuals, which may limit the ability to fully disentangle temporal and cohort effects. Second, aggregation of individual-level data may reduce heterogeneity and introduce ecological bias. Finally, as with all observational data, the identified associations should be interpreted as predictive rather than causal.

Despite these limitations, our study demonstrates the feasibility and value of combining sero-prevalence cohort data with routine surveillance data for incidence prediction. The consistency of key predictors across multiple model classes and explainability methods supports the robustness of the observed patterns, while differences between time-aware and non-temporal models, as well as between SHAP and LIME under differential privacy, highlight how methodological choices influence the representation and interpretation of these signals. From a digital health perspective, these results support the integration of cohort-based data streams into surveillance and decision-support systems under appropriate privacy-preserving mechanisms, and provide a foundation for extending such approaches with spatial information, richer temporal representations, and alternative privacy-preserving techniques.

## Data Availability

The data underlying this study are not publicly available due to privacy and ethical restrictions related to individual-level health data. Access can be requested through the MuSPAD data access process via https://serohub.net/, subject to approval by the data custodians and applicable data protection regulations. The code used for the analyses, as well as the data codebooks and top-50 explainability tables are available at https://github.com/JesKre/Using-machine-learning-on-seroprevalence-studies-to-obtain-predictors-for-COVID-19-incidence_code

## Data and code availability statement

The data underlying this study are not publicly available due to privacy and ethical restrictions related to individual-level health data. Access can be requested through the MuSPAD data access process via https://serohub.net/, subject to approval by the data custodians and applicable data protection regulations. The code used for the analyses, as well as the data codebooks and top-50 explainability tables are available at https://github.com/JesKre/Using-machine-learning-on-seroprevalencecode.

## Acknowledgments

This work was supported by the Initiative and Networking Fund of the Helmholtz Association, Germany (grant agreement number KA1-Co-08, Project LOKI-Pandemics).

## Supplementary Material

### S1 Privacy Implementation Details

#### Definition 1

(Differential Privacy [48]). *A randomized mechanism M with range R satisfies* (*ε, δ*)*-differential privacy, if for any two adjacent datasets E and E*^*′*^, *i*.*e*., *E*^*′*^ = *E ∪ {x} for some x in the data domain (or vice versa), and for any subset of outputs O ⊆ R, it holds that*

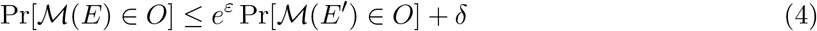

Intuitively, differential privacy (DP) guarantees that an adversary, given the output of *ℳ*, cannot draw significantly different conclusions (up to *ε* with probability larger than 1 *™ δ*) about any record, regardless of whether it is included in the input of *ℳ* or not [48]. This means that, for any record owner, a privacy breach is unlikely to occur due to their participation in the dataset.

#### S1.1 Implementation details

In our study, we address privacy concerns by employing Differentially Private Stochastic Gradient Descent [49] (DP-SGD) while training an ML model. The procedure begins by randomly selecting a batch of data using Poisson subsampling, which ensures that each data point is included in the batch with a fixed probability, independent of others. This reduces the number of data points seen during each update. Next, we clip the norm of the gradients to constrain their sensitivity, ensuring that no individual data point can disproportionately influence the model’s updates. Finally, we add carefully calibrated noise to the aggregated clipped gradients, which ensures that the gradients do not reveal sensitive information about individual records. This process prevents the model from overfitting to individual data points, thereby reducing the risk of memorization or leakage of sensitive details. By ensuring that the model learns general patterns rather than specific records, DP-SGD aligns with the principles of DP and provides strong theoretical guarantees against privacy breaches.

We implemented the DP-SGD approach in R, as existing libraries for DP-SGD are primarily available in languages like Python (e.g., Opacus [56]). Our implementation follows the methodology described in the original DP-SGD paper [49], adapting it to the R ecosystem. Additionally, we quantified the privacy guarantees in the domain of Rényi Differential Privacy (RDP), which provides a more precise framework for analyzing privacy loss under composition. We then converted the RDP privacy parameters to the (*ε, δ*)-DP domain.

**Table S1:**
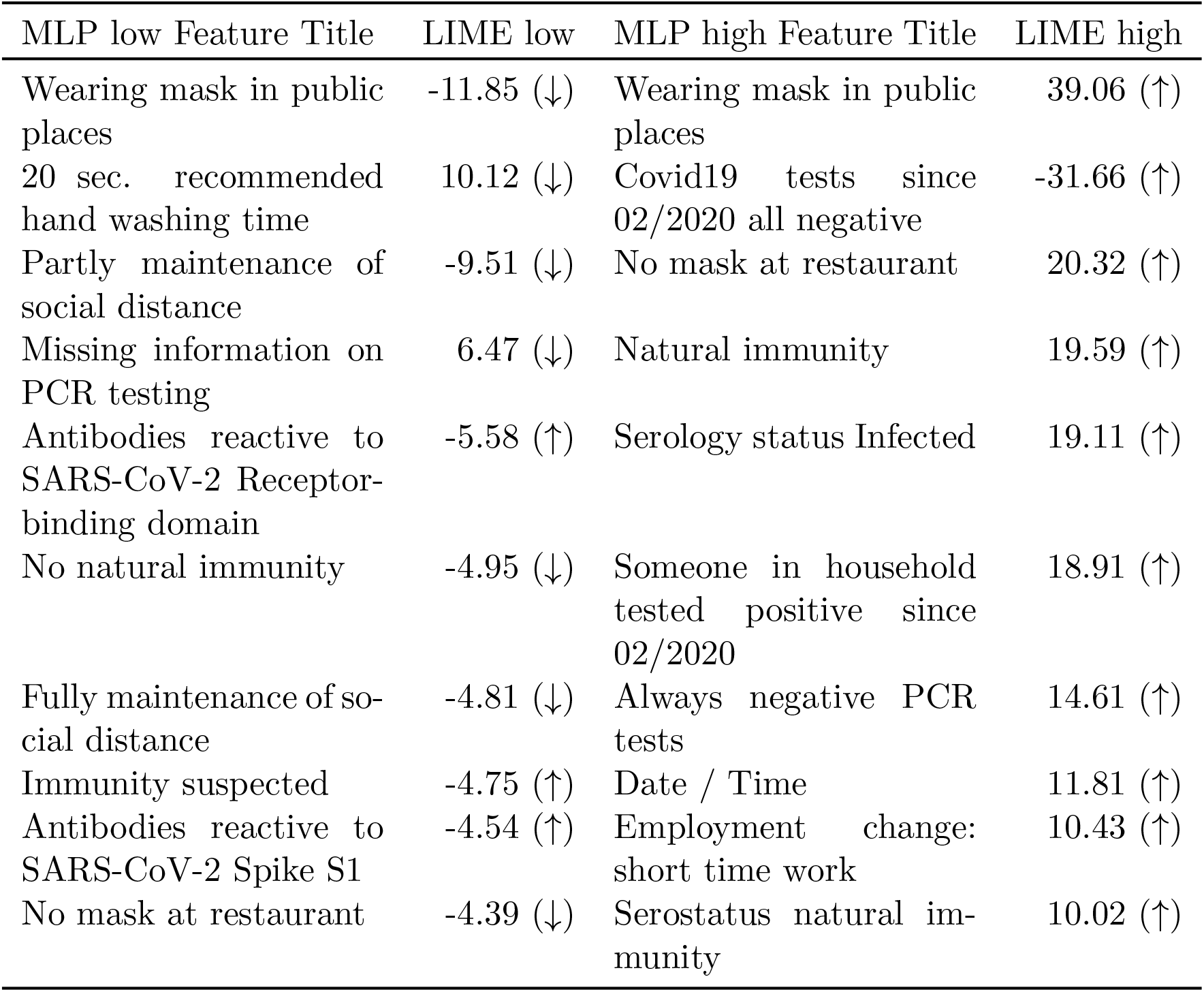
Explainability table of LIME values for the 10 most important features (ranked by absolute value) in low-and high-incidence clusters for the MLP model. “MLP low Feature Title” and “LIME low” contain information on low incidence cluster; “MLP high Feature Title” and “LIME high” contain information on high incidence cluster. (↓) indicates that mean of feature values in cluster (feature cluster mean) is below overall mean of feature values (feature overall mean); (↑) indicates that feature cluster mean is above feature overall mean.

**Table S2:**
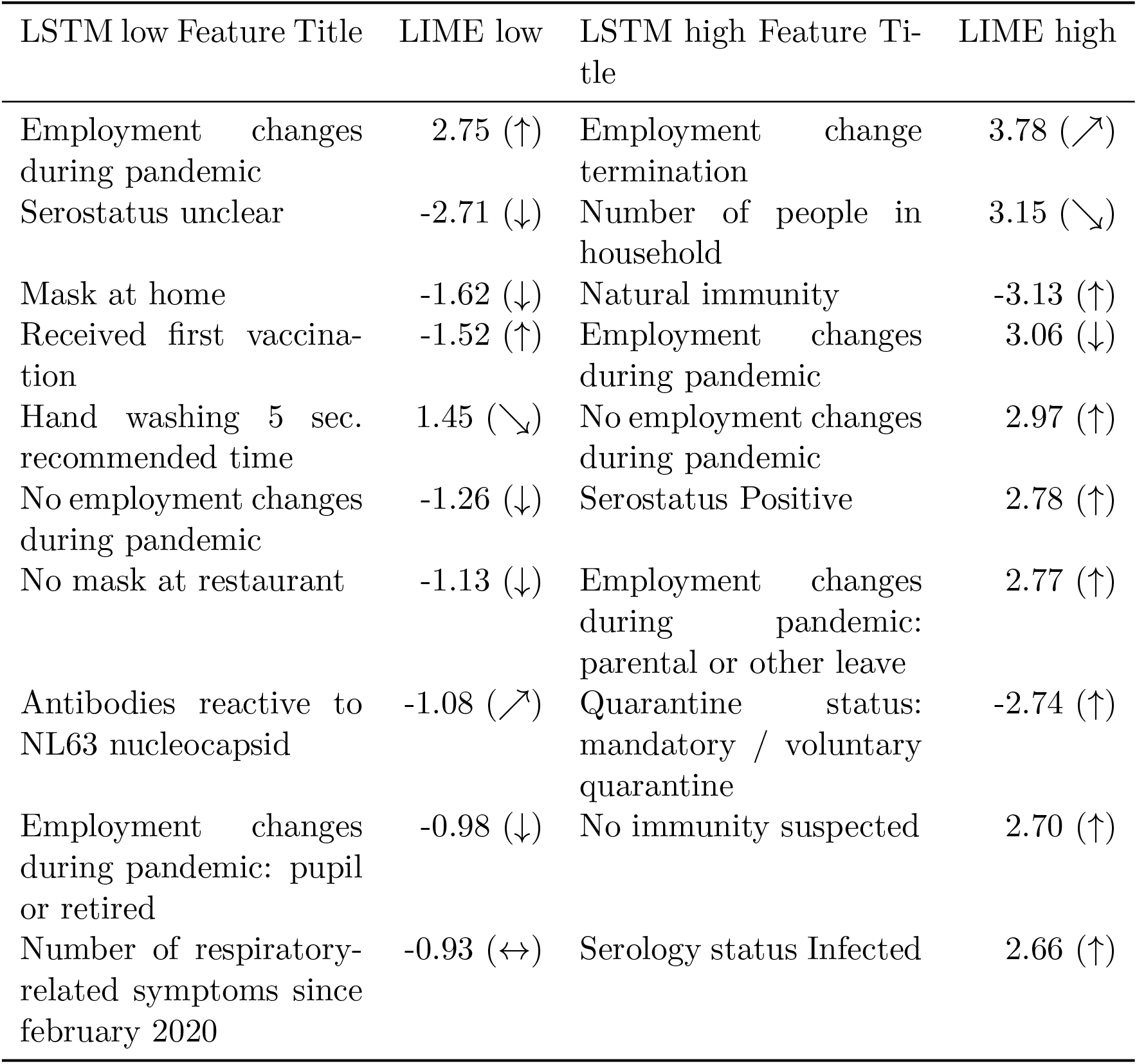
Explainability table of LIME values for the 10 most important features (ranked by absolute value) in low-and high-incidence clusters for the LSTM model. (*↘*) indicates that feature cluster mean is less than 0.06 below feature overall mean, (*↗*) indicates that feature cluster mean is less than 0.06 above feature overall mean. The remaining notation is as in Table S1.

## Notes

### Competing Interest Statement

The authors have declared no competing interest.

### Author Declarations

MuSPAD complies with all relevant laws and declarations (EU Charter of Fundamental Rights, Biomedical Convention of the Council of Europe and additional protocols, the CIOMS guidelines and the Helsinki Declaration), ethical approval was given on 21.06.2020 by the Ethics Committee of the Hannover Medical School (Ethics approval no 9086_BO_S_2020). All necessary patient/participant consent has been obtained and the appropriate institutional forms have been archived.

